# Women’s Experiences of Accessing and Using Patient Portals Across Health Settings and Implications for Mental Health Care: A Qualitative Descriptive Study

**DOI:** 10.64898/2026.05.03.26352340

**Authors:** Keri Durocher, Jessica Kemp, Hwayeon Danielle Shin, Kimberley T. Jackson, Gillian Strudwick

## Abstract

Patient portals are online tools that enhance patients’ access to various aspects of their health care, including provider communication, medication information, and lab results. As portals continue to be integrated into health systems, it is imperative to understand the experiences of various groups who utilize their functions. Women’s experiences of using patient portals have been scantly explored in the literature, including their perceptions about use for mental health care. The purpose of this study was to explore women’s experiences of accessing and using a variety of patient portals, including their perceptions of usefulness for mental health care. A qualitative descriptive methodology was used to explore women’s experiences of accessing and using patient portals across Canada. Purposive sampling was used to recruit ten women, who completed semi-structured, one-to-one interviews between April-June 2025. Conventional qualitative content analysis was used to analyze the data. Each woman had used at least one patient portal for their health care at the time of their interview. Four main themes emerged from the data, including: (1) the health care lived experience, (2) individual autonomy, (3) provider partnership, and (4) portal improvement. The interrelated themes contain narrative descriptions of individual experiences of accessing and using patient portals, and implications for using portals for women’s mental health care. These results demonstrate a variety of women’s experiences. Patient portals were found to impact their lived experiences with health care, enhance individual autonomy, and foster partnerships with their health care providers. The women also suggested various areas of improvement in portal design elements, features, and privacy functions. Future research should focus on evaluating the design of new portals to ensure they meet the needs of the population they serve.

**Author Summary:** A patient portal is an example of a digital tool that is being integrated into various health organizations to supplement in-person care. Depending on the design and the complexity of the portal, patients may be able to complete online prescription renewals, access medication schedules, virtually communicate with their providers, and review their clinical notes. However, as digital tools continue to be produced and adapted within health settings, it is crucial to understand how they can best serve different populations. In this study, we explored women’s experiences with using patient portals for their health care in Canada. We also aimed to understand women’s perspectives on how patient portal use can be optimized for mental health care. We performed virtual interviews with 10 women who had used at least one patient portal for their health care, and gained their perspectives on accessibility, useful features, and how using a patient portal impacted their experiences of receiving health care. The women discussed how portal use improved their health care experiences and they suggested a variety of features to support mental health care as patient portal designs continue to be adapted to different settings.

## 1. BACKGROUND

A patient portal is a secure, online tool that allows patients to access various aspects of their health care in a digital format, including electronic personal health records, secure messaging with health care providers, lab results, and medication lists.(1,2) Improvements in the functionalities of patient portals are continuously being advanced to increase health care access for various patient populations. For example, recent systematic review findings detailed how the delivery of education materials through a patient portal enhances the utilization of these resources, leading to better health outcomes.(3) Additionally, a recent cross-sectional survey detailed how positive experiences with patient portals were linked to specific functions within the portal, such as prescription renewals.(4) Identifying the specific functions that are related to positive user experiences can be beneficial when prioritizing functionalities during portal implementation.

As the development and use of patient portals continues to evolve, it is imperative to understand experiences amongst the populations who access them to optimize the benefits of using patient portals. Women may be more likely to engage with digital health tools when compared to men(5) however, their personal experiences with accessing and using patient portals for their health care has been scantly explored within the literature. We recently published a rapid review about this phenomenon which included 15 articles, and explored the uptake and use of patient portals for women in a variety of settings and with different conditions.(6) For example, patient portals are now frequently being integrated into obstetric and gynecological settings(7,8), with specific features such as appointment reminders, informational videos, and medication management systems.(9)

Additionally, no studies to date have included women’s individual experiences and perceptions of using a patient portal for mental health care. Other specific populations have been investigated throughout the literature, such as in a review conducted by Rabbani et al. (2023), which provided general recommendations for patient portal features for mental health care including direct provider messaging and medication refill options.(10) Hornum et al. (2023) explored adolescents’ experiences with using a mental health patient portal and provided direct recommendations for portal use within these settings to guide providers, parents, and patients in optimal use.(11) Recently, a large German cohort study presented findings that showed how women and men differ in both risk and protective factors for mental health.(12) For example, women consistently self-reported worse mental health than men and had higher rates of depression, anxiety, post-traumatic stress, and suicidal ideation.(12) Women can also encounter sex-specific barriers in accessing mental health care services within different health contexts(13,14); therefore, understanding their experiences and perceptions of using a patient portal for mental health care is imperative to enhancing future use. For example, Sambrook Smith et al. (2019) conducted a systematic review and discovered that within the perinatal period, women encounter numerous barriers to access mental health resources, including unclear policies, stigma, and health literacy implications.(13) Secondly, a recent qualitative study highlighted the structural barriers that women experience in accessing victim support after facing intimate partner violence, specifically related to mental health support.(14)

In this qualitative descriptive study, we aimed to describe the experiences of women in accessing and using a variety of patient portals across Canada, including their perceptions of usefulness and relevance to mental health care. In health research, qualitative description is a design used to thoroughly understand patient experiences related to their health care.(15) This study is the second phase of a larger project, where we are generating actionable recommendations for improvement to the patient portal for the largest mental health organization in Canada.

## 3. METHODS

### Research Design and Sampling Techniques

Qualitative description is a methodology that enhances the discovery of insightful information about the experiences of individuals who shared a similar phenomenon.(16,17) This methodology draws from the grand theory of naturalistic inquiry, which seeks to examine specific individual experiences within a natural setting.(16,18) This methodology is flexible, which means that some of the described methods may have overtones from other methodological traditions. Descriptions, as provided by the participants, will vary based on their values, experiences, and interpretations.(16) Within this study, the central phenomenon that participants experienced was accessing a patient portal for services related to women’s health, including their perceptions of portal use for mental health care.

The inclusion criteria for this study involved any woman who had accessed a patient portal within Canada for health care services (including mental health care). Participants also had to have access to a device (laptop, smartphone, etc.) for the virtual interview and verbal communication skills in English. Purposive sampling was used to recruit participants through the following methods: (1) a virtual ad posted within the study setting’s patient portal, social media sites, and sent out through relevant networks, and (2) hardcopy posters placed in common areas at the study setting.

### Data Collection

Ten women completed semi-structured, virtual interviews that took place via Webex videoconferencing between April-June 2025. Interested, potential participants contacted the research team by email, where their eligibility for participation was assessed. We did not collect demographic data due to the small sample size and risk of identification. If eligible for enrolment, potential participants received a copy of the informed consent form to review and scheduled a consent review session with KD as desired. After signing the informed consent form, eligible participants were enrolled into the study. The research team signed the form and sent the participant the completed copy. All interviews were between 15-60 minutes in length, and safeguarding activities were implemented to maintain participant confidentiality, including password-protection and a waiting room for the videoconferencing session. In thanks for their participation, all participants were offered an honorarium in the form of a $25 electronic gift card. The semi-structured questions focused on the following topics: (1) each woman’s individual experiences with patient portals, (2) improvements they would make to the portal(s) they have accessed, and (3) their perceptions of using patient portals for women’s mental health care. To access the full interview guide, please see Table 1.

**Table 1.** Interview Guide.

**Interview Guide**
| Topics | Questions |
| --- | --- |
| Introduction questions | • How do you feel about the use of a patient portal?<br>• Why did you decide to access the patient portal?<br>• How often do you access the patient portal? |
| Opinions and experiences with patient portals | • What do you like about the patient portal?<br>• What do you not like about the patient portal?<br>• What features of the patient portal do you find helpful?<br>• What features of the patient portal do you find not helpful?<br>• Did using the patient portal impact your experience of receiving health care? |
| Patient portals and women's mental health care | • Do you think the patient portal is beneficial for women's mental health care?<br>• Do you feel that women have any barriers to accessing the patient portal?<br>• Can you think of any features that could be added to the patient portal that would be beneficial for women's mental health care? |
| Conclusion questions | • Is there anything else that you would like to share about using the patient portal?<br>• Would you like us to follow up with you on any recommendations that have been made based on these interviews? |

### Data Analysis

Conventional qualitative content analysis was used to analyze the qualitative data. This method is one of the most common analysis approaches within qualitative descriptive studies.(16) This approach promotes the processes of staying close to the collected data and summarizing the contents, which are both integral to understanding a specific shared experience. Prior to initiating coding, key words were highlighted to identify important concepts and to assist with data immersion.(19) Borrowing from a grounded theory methodology, open coding (an inductive approach) was performed for each interview transcript.(16) The primary author (KD) coded all the data, and two members of the research team (JK, HDS) verified the coding procedures. NVivo software was used to assist with data analysis.(20) From specific codes, broader categories and sub-categories were formed, and relationships between them were investigated. It is important to note that the analysis went beyond simply listing what was said by the participants but was used to interpret connections and the importance of these findings.

## 4. RESULTS

The women had used at least one patient portal for their health care in Canada at the time of the interviews. Some women had used a portal for their mental health care, and all were asked to share their perspectives on use of a portal for mental health. The women were from a variety of provinces across the country and had accessed portals across various levels of health care, from primary to tertiary. Four key themes emerged from the data analysis procedures, including: (1) the health care lived experience, (2) individual autonomy, (3) provider partnership, and (4) portal improvement. Each theme had a variety of corresponding categories that developed from iterative coding procedures. For a full list of themes and category definitions, please refer to Table 2.

**Table 2.** Data Analysis Structure.

| Theme | Theme Definition | Categories | Category Definition |
| --- | --- | --- | --- |
| The Health Care Lived Experience | Women's personal experiences of using patient portal(s) for various aspects of their health care. | 1. Accessibility<br>2. Convenience<br>3. Comprehensiveness | 1. How the availability of portals and their features can enhance women's health care.<br>2. The ease of use of patient portals for women's health care.<br>3. The inclusiveness of a variety of features within patient portals. |
| Individual Autonomy | How accessing and utilizing patient portal(s) can enhance women's autonomy for their health care. | 1. Empowerment<br>2. Engagement<br>3. Transparency<br>4. Awareness<br>5. Recall | 1. How use of patient portal(s) can provide a sense of power for women.<br>2. How women's involvement and interest in their health care can be improved through use of patient portal(s).<br>3. The importance of honesty and openness in health care that can be facilitated using portal(s).<br>4. Women's understanding and comprehension of their health care.<br>5. How portal(s) can help women remember various aspects of their health care. |
| Provider Partnership | How portal use between health care providers and women can build relationships and enhance health care access. | 1. Interprofessional Collaboration<br>2. Continuity of Care | 1. Relationships amongst providers and patients for health care decision making.<br>2. Ensuring that women's health care is coordinated between providers. |
| Portal Improvement | How portals can be improved to increase women's access and utilization. | 1. Portal Design<br>2. Portal Features<br>3. Privacy | 1. Women's perspectives about the design elements of a variety of portals.<br>2. Women's perspectives about current or needed features to improve portals.<br>3. Confidentiality considerations for portal use. |

### The Health Care Lived Experience

The personal experiences of using patient portals were described by each woman in relation to various parts of their health care. From these lived experiences, accessibility through a variety of portal features was described as a key important factor. Many women described how the availability of specific features and design elements enhanced their health care experiences. Some of these elements included “simple language” (Participant 10) or features such as “appointment booking” (Participant 7), engaging in “group therapy” sessions (Participant 4) and the ability to access “consent forms” (Participant 6). One participant discussed how accessibility can enhance safety for some women:

*One thing that I can think of that’s a potential benefit for women is the communication with the care team because it’s kind of like communications that are not stored on your computer. They’re not part of your email history. So, if you were trying to communicate with staff, and you do not want your partner to know what you were saying, then that I think could be a real plus if, for example, say a woman was in an abusive relationship.* (Participant 4)

Many participants described how using patient portals enhances convenience for women, specifically in the time saved “travelling” (Participant 10) while managing other responsibilities and having “quick access” to medical records (Participant 9). They can also be helpful for managing appointments, such as “psychiatry referrals” (Participant 1) or for “looking at biographies of therapists” via specific search functions (Participant 6). One participant described the process of how she previously booked appointments versus how using a patient portal has saved her time:

*“I used to call and it would ring and it would ring, and it would ring and you’re put on hold. Then, you know, nobody would answer, then you’d have to try and it would take sometimes two or three hours of your time to call. Now you just go online, go to the portal, OK I’ll take that appointment, book it and it’s done.”* (Participant 7)

Women also described how the comprehensiveness of portals can enhance their user experience, as the inclusion of a variety of features helps with health care navigation. A few women described some improvements that could be made to the portals they have used, including “education” elements for women’s health (Participant 2), and having access to “lab results, imaging and records” (Participant 5). Other participants echoed the need for education elements including having access to “courses and resources” (Participant 8) and “self-paced education modules” (Participant 6).

From a mental health perspective, having access to mental health history records may be helpful in a central portal. As described by one participant, details of past mental health interventions can show future providers how to guide further care interventions: *I was always kind of worried about sharing my mental health status or the drugs I have been prescribed because I didn’t really have a family doctor here. So, I was always a little skeptical when it came to disclosing that information. But when I looked back on this record, I saw that I had disclosed this to the nurse at the ER. So, I was like OK, if I would have to go back for whatever reason to show that this is what my history is, I do have some kind of proof to say that yes, even then I did disclose it, and I have been on this medication since so-and-so date.* (Participant 5)

### Individual Autonomy

The women described significant increases in their individual autonomy for managing their health care through the utilization and access of patient portals. Many women described a sense of empowerment that comes through portal use. Having access to clinical notes can foster self “advocacy” (Participant 3). Self-initiation of portal use can else enhance women’s sense of power:

*Some people will only do things at their own discretion as opposed to somebody else pushing them to do it. So that could be very much self-empowering to be able to say I’m doing this, booking it on my own. You know? Like one of those things. Yeah. So, people could feel empowered by booking it themselves.* (Participant 8)

Through the women’s descriptions, engagement with their health care was also reported as improved through the integration of patient portals. One woman reported increased interest due to “curiosity” about her health care (Participant 8) while another described that she is more engaged with her health care due to the portal making management “easier” (Participant 10). One participant described how having access to her diagnostic imaging results increased engagement with her provider:

*I didn’t have to just rely on doctors to tell me what happened and, you know, I could actually look at their records. And, you know, like there were sometimes I would see something written about the x-ray that they didn’t share with me and then I would bring it up at the next appointment.* (Participant 9)

Women also described how using patient portals enhanced the transparency of their individual health care, specifically the facilitation of honesty and openness between the women and their providers. This transparency can assist with women being “involved in care” (Participant 1) and having access to “all of the information” (Participant 9). One woman described the dangers of not having access to her health care information, and how this can interrupt continuity of care:

*I’m with a gynecologist and a menopause doctor. And they currently don’t put any notes, any imaging, nothing online. And they actually don’t send anything to my family doctor. So, nothing is transparent there. And for someone in complex medical conditions, there’s a couple barriers there.* (Participant 8)

Awareness was another factor that could be enhanced by patient portals, including women’s understanding and comprehension of their health care. One woman commented how she “appreciated” (Participant 1) being able to read medical notes from her family doctor due to confusion after speaking to her psychiatrist. Some women suggested areas of improvement that could enhance awareness, including “incorporating an AI feature” for translation of medical jargon (Participant 3) and having “reminders” in the system for appointments (Participant 8). One woman described how having quick access to lab results assists with her awareness of her condition:

*I’m on a lot of different medications to treat my disease; they can affect the kidneys; they can affect liver function. So, I can go and check. When I go on and get my blood work, I can go that night sometimes like after – if I have it in the morning, they’ll be done – they will be able to post it by the evening or the next day and so I can compare to the ones I had three months previous.* (Participant 7)

Lastly, the women described how patient portals could help with recall, defined as remembering various aspects of their health care. One woman appreciated the “chronological timeline” (Participant 3) of her health care data within the portal while another stated that a “reminder system” for appointments would be helpful (Participant 8). For mental health care, one woman described how having access to certain statistics could assist in recovery:

*For someone let’s say who has mental health issues or for some reason they’re not able to attend their appointments, I think it would serve greatly to that person because it would serve themselves like a reminder that these are the messages that they’ve had.* (Participant 10)

Another woman described how individual autonomy of managing one’s mental health care could be improved with a comprehensive, accessible patient portal:

*When you have something else in the future or some other thing that you’re trying to decide what to do like with your care or your mental health, to be able to log into one place and be able to access. Because I tried to remember my history and be like you know when did I start taking that? And when did I need to increase it and what had happened at that point in my life? Or what did I do to get better when it had to go down? So, if I could really track my own life, it would be good for my own planning.* (Participant 1)

### Provider Partnership

The women described how using a patient portal in collaboration with their health care providers built strong relationships to enhance access to care services. Relationships were built through interprofessional collaboration between providers and patients, to enhance autonomy in decision making. This was sometimes facilitated through the “messaging function” (Participant 3) and “Zoom appointments” (Participant 4). One woman described her enhanced understanding of her medical care by viewing her medical notes through a portal:

*I had, you know, a better idea of what was going on, and I could kind of address. Like I didn’t have to just rely on doctors to tell me what happened and, you know, I could actually look at their records.* (Participant 9)

In addition to collaborating with providers, the women described how having access to a patient portal led to a greater continuity of care through coordination between providers. This continuity was believed to create greater “transparency” (Participant 1) through greater patient involvement and “streamline” (Participant 4) some administrative procedures. One woman described how requesting past medical records through the portal could assist with continuity amongst various health care providers:

*And if it’s a new doctor, or you’ve had diagnoses like I have for the past 25 years. So, that’s what I had to do. I had to find all these records, and it was very challenging, and also an expensive process because they usually like to charge you for those. But even if I could just request through the portal, access some of those records, I think that would really take a lot of the burden off.* (Participant 6)

To continue to promote provider partnership, the participants suggested further improvements that can be made to enhance health care access. Providing a “directory” (Participant 8) of health care providers and specialized “alerts” (Participant 7) for general providers in the case of emergencies are two innovative ways to enhance collaborative care efforts. Additionally, women could utilize patient portals to keep track of some health activities, including diagnostic tests and vaccination schedules:

*Maybe you could have something in the portal that tells you that this is what the eligibility criteria for this test is. If you qualify, you should get a test. Or this is the time that has elapsed since you last got your vaccine or immunization. It’s now time for you to go get it. Even for something like breast cancer screening.* (Participant 5)

### Portal Improvement

Throughout the interviews, the women consistently described how elements of patient portals can be improved to increase women’s access and utilization. Specifically, the design of portals can impact the user experience depending on the “website provider” (Participant 2) and the “format or layout” (Participant 3). One woman described a key consideration for organizations to consider when implementing their portal design:

*Sometimes the patient portals have different texts. So, for example somebody who is neurodivergent, I think it’s very difficult to just keep – your attention just goes here and there and you’re not able to focus because there’s so much variety in text on the portal. So, I feel that is something that distracts me. So, font size, font color, one size of text.* (Participant 10)

The features that portals offer was also viewed as highly important. Various favorable features were mentioned including “artificial intelligence” (Participant 3) for translating medical jargon, “communication [features] with your care team” (Participant 3) and having a “question and answer section” (Participant 4) for understanding how to navigate the portal. Regarding mental health and trauma support through group therapy, seeing “what courses are available and when and to sign up” (Participant 8) was also described as helpful. One woman described a change that could help enhance women’s understanding of their health care:

*I think it could be as simple as click for a fact sheet. I think we need to always consider, now, I’m big in the patient engagement space, and simple plain language, right? Maybe it’s a brief description of what this test is for. It’s like they’re named things like Biolase Immunace, and I’m like, what is this? So, just a brief description of what it’s for, a brief description of the healthy range, and what it can mean to fall outside of the range in either way.* (Participant 6)

Confidentiality and privacy considerations when using patient portals were commonly described by the women. With the increased use of technology for health care, the women described specific concerns including “phishing” (Participant 10) and having “certain sections password protected” (Participant 5). A few participants described some of the accessibility issues that women may have with using a portal, and why privacy is such an important consideration. As described by one woman:

*“Keeping intersectionality in mind, I was just thinking about women that might have not access to gadgets or might not feel comfortable accessing these digital platforms on their phones or on shared devices. [They] might not want to share certain information with other people in their families or like among loved ones.”* (Participant 5)

## 5. DISCUSSION

In this qualitative descriptive study, we explored women’s experiences of accessing and using patient portals for their health care within Canada, including their experiences and recommendations for using these portals for mental health care. The results demonstrate how patient portals can impact a woman’s lived experience of her health care, enhance individual autonomy, and foster connections with health care providers that go beyond in-person appointments. Various improvements to patient portals were also discussed. From a mental health perspective, Kipping et al. (2016) discovered significant improvements in recovery domains with patient portal use(21). Therefore, to support women’s mental health, it is imperative to understand women’s perspectives on patient portal use for this area of health care.

Within the findings, the women discussed how accessing a patient portal enhanced accessibility to their health care, improved convenience, and fostered comprehensiveness when a variety of features were included. A recent mixed methods study on patient portal use related to functioning and recovery in a mental health setting also showed qualitative results where patients related portal use to enhanced convenience for accessing their health care(22). Another study examined gender differences in accessing patient portals in Northern Europe and concluded that women preferred more functionalities and comprehensive access to information when compared to men(5). These findings echo the importance of having user-friendly features when designing portals, which can enhance accessibility.

Our study results demonstrate how accessing and utilizing patient portals can enhance individual autonomy for women when managing their health care through improvements in empowerment and engagement with their health, improving transparency and awareness of their health care, and assisting with recall. Other studies have included results that demonstrate how frequent engagement with a patient portal has improved health outcomes and treatment compliance. For example, Zhang et al. (2024) showed positive weight loss outcomes after bariatric surgery when patients had higher engagement levels with a patient portal(23). Szukagyi et al. (2020) demonstrated an increase in influenza vaccination rates by providing reminders and education materials to patients via a patient portal(24). A recent scoping review also highlighted certain success factors in relation to mental health care, including availability of resources and educational materials that can contribute to enhanced autonomy for this population(25).

Provider partnership was explored in this study and examined how interprofessional collaboration and continuity of care can be fostered through portal use to enhance these relationships for women. Two recent mixed methods studies explored compassion(26) and interprofessional collaboration(27) for mental health patients who accessed a patient portal and found that enhancements can be made through effective portal use in mental health care settings. Recent systematic review findings also revealed that improvements can be made through provider-patient interactions and perceived enhancements in care when using a patient portal(28). Lastly, various ideas for portal improvement were shared by the women, including enhancements to design, features, and privacy settings. Most notably, the need for a wide variety of features was discussed to enhance usability. Recent systematic review findings have revealed that there are almost 200 features across a variety of portals that are currently in use(29). Therefore, it is imperative that health care organizations understand the features that are most helpful for the populations they serve.

## Limitations

To our knowledge, this is the first study to explicitly explore women’s experiences of using patient portals for their health care, in addition to their perspectives on portal use for supporting mental health. Our recent rapid review(6) revealed how women’s mental health care has not been examined in relation to patient portal use, and future research can gather a wider variety of perspectives from women with mental health challenges or that are connected to specific mental health organizations to implement tailored patient portal designs. Secondly, we omitted gathering demographic data due to the small sample size and risk for identification. Future research can explore portal use alongside specific demographic variables to further explore how to best meet the needs of varying populations.

## 6. CONCLUSIONS

The results of this qualitative descriptive study demonstrate a variety of women’s experiences of using patient portals for their health needs, including their perspectives and experiences with utilizing them for mental health care. For women, patient portals were found to impact their lived experiences with health care, enhance their individual autonomy, and foster partnerships with their health care providers. The women also suggested various areas of improvement in portal design elements, including features, and privacy functions. Future research should focus on evaluating the design of new portals that are tailored to specific populations to ensure the needs of various groups are being met, including for women’s health. As the complexity and design of portals continues to evolve, focusing on these key elements may help women have better mental health outcomes, and overall access to health care.

## Data Availability

The datasets used and/or analyzed during the current study are available from the primary author on reasonable request.

## 7. LIST OF ABBREVIATIONS

N/A

## 8. DECLARATIONS

### 8.1 Ethics approval and consent to participate

Ethical approval for human participation was obtained from the Research Ethics Board at the Centre for Addiction and Mental Health (Reference Number 2023/207).

### 8.2 Consent for publication

Not applicable

### 8.3 Availability of data and materials

The datasets used and/or analyzed during the current study are available from the corresponding author on reasonable request.

### 8.4 Competing Interests

The authors declare that they have no competing interests.

### 8.5 Financial Disclosure Statement

This work was supported by the Health System Impact Fellowship (PhD) from the Canadian Institutes of Health Research. The funders had no role in study design, data collection and analysis, decision to publish, or preparation of the manuscript.

### 8.5 Authors’ contributions

The content of this manuscript was conceptualized by KD in collaboration with all authors. KD was responsible for conducting the participant interviews, drafting the original manuscript and overseeing revisions by the other authors. KD, HDS and JK performed all data analysis procedures. GS and KTJ were KD’s doctoral supervisors during her fellowship. KD, GS, HDS, JK, and KTJ were thoroughly involved in revising the manuscript prior to submitting it for publication consideration. All authors agree regarding the final version of the manuscript.

## 8.6 Acknowledgements

Not applicable

## Notes

### Competing Interest Statement

The authors have declared no competing interest.

## REFERENCES

1. Assistant Secretary for Technology Policy. What is a patient portal? [Internet]. HealthIT.gov; 2025. Available from: https://www.healthit.gov/faq/what-patient-portal

2. Ontario Health. Patient portal standards [Internet]. 2025. Available from: https://www.ontariohealth.ca/digital/standards/patient-portals

3. Johnson AM, Brimhall AS, Johnson ET, Hodgson J, Didericksen K, Pye J, et al. A systematic review of the effectiveness of patient education through patient portals. JAMIA Open. 2023;6(1):ooac085. doi:10.1093/jamiaopen/ooac085

4. Simola S, Hörhammer I, Xu Y, Bärkås A, Fagerlund AJ, Hagström J, et al. Patients’ experiences of a national patient portal and its usability: cross-sectional survey study. J Med Internet Res. 2023;25:e45974. doi:10.2196/45974

5. Kharko A, Hagström J, Simola S, Cajander A, Blease C, Hägglund M. The gender gap in EHR experiences and preferences: results from an international cross-sectional survey of patients. Stud Health Technol Inform. 2025;(327):939–43. doi:10.3233/SHTI250510

6. Durocher K, Shin HD, Jackson KT, Strudwick G. Women’s experiences of using patient portals in healthcare settings: a rapid review. BMC Womens Health. 2024;24(449). doi:10.1186/s12905-024-03292-9

7. Kalejta CD, Higgins S, Kershberg H, Greenberg J, Alvarado M, Cooke K, et al. Evaluation of an automated process for disclosure of negative noninvasive prenatal test results. J Genet Couns. 2019;28(4):847–55. doi:10.1002/jgc4.1127

8. Forster M, Dennison K, Callen J, Georgiou A, Westbrook JI. Maternity patients’ access to their electronic medical records: use and perspectives of a patient portal. Health Inf Manag J. 2015;44(1):4–11. doi:10.1177/183335831504400101

9. MacEwan SR, Fareed N, Jonnalagadda P, Heffer H, Petrecca AM, McAlearney A. Patient and provider perspectives on the use of patient portals during pregnancy and the postpartum period. J Telemed Telecare. 2023;31(2):277–85. 10.1177/1357633X231177742

10. Rabbani M, Nasiri M, Mowla A, Sharifian R. Mental health patient portals aimed at depression: a picture close to reality. Stud Health Technol Inform. 2023;(311):45–53. doi:10.3233/SHTI200608

11. Hornum MS, Steinsbekk A, Nøst T. Views on patient portal use for adolescents in mental health care - a qualitative study. BMC Health Serv Res. 2023;23(132). doi:10.1186/s12913-023-09156-6

12. Otten D, Tibubos AN, Schomerus G, Brähler E, Binder H, Kruse J, et al. Similarities and differences of mental health in women and men: A systematic review of findings in three large German cohorts. Front Public Health. 2021;9. doi:10.3389/fpubh.2021.553071

13. Sambrook Smith M, Lawrence V, Sadler E, Easter A. Barriers to accessing mental health services for women with perinatal mental illness: systematic review and meta-synthesis of qualitative studies in the UK. BMJ Open. 2019;9:e024803. doi:10.1136/bmjopen-2018-024803

14. Daoud M, Saleh-Darawshy NA. Multiple barriers for accessing mental health service among women attending shelters for women experiencing intimate partner violence. Eur Psychiatry. 2022;65:S320. 10.1192/j.eurpsy.2022.815

15. Doyle L, McCabe C, Keogh B, Brady A, McCann M. An overview of the qualitative descriptive design within nursing research. J Res Nurs. 25(5):443–55. 10.1177/1744987119880234

16. Sandelowski M. Whatever happened to qualitative description? Res Nurs Health. 2000;23(4):334–40. 10.1002/1098-240x(200008)23:4%3C334::aid-nur9%3E3.0.co;2-g

17. Neergaard MA, Olesen F, Andersen RS, Sondergaard J. Qualitative descripion - the poor cousin of health research? BMC Med Res Methodol. 2009;9(52). doi:10.1186/1471-2288-9-52

18. Sandelowski M. What’s in a name? Qualitative description revisited. Res Nurs Health. 2010;33(1):77–84. doi:10.1002/nur.20362

19. Guest G, Namey E. A simple method to assess and report thematic saturation in qualitative research. PLOS One. 2020;15(5):e0232076. doi:10.1371/journal.pone.0232076

20. Lumivero. NVivo [Internet]. 2026. Available from: https://lumivero.com/products/nvivo/

21. Kipping S, Stuckey MI, Hernandez A, Nguyen T, Riahi S. A web-based patient portal for mental health care: Benefits evaluation. J Med Internet Res. 2016;18(11):e294. doi:10.2196/jmir.6483

22. Lo B, Shin H, Kemp J, Munnery M, Chen S, Ma C, et al. Shifting mindsets: The impact of a patient portal on functioning and recovery in a mental health setting. Can J Psychiatry. 2023;69(3):217–27. doi:10.1177/07067437231197060

23. Zhang X, Kang K, Yan C, Feng Y, Vandekar S, Yu D, et al. Association between patient portal engagement and weight loss outcomes in patients after bariatric surgery: Longitudinal observational study using electronic health records. J Med Internet Res. 2024;26:e56573. doi:10.2196/56573

24. Szilagyi P, Albertin C, Casillas A. Effect of patient portal reminders sent by a health care system on influenza vaccination rates. JAMA Intern Med. 2020;180(7):962–70. doi:10.1001/jamainternmed.2020.1602

25. Zhang T, Shen N, Booth R, LaChance J, Jackson B, Strudwick G. Supporting the use of patient portals in mental health settings: A scoping review. Inform Health Soc Care. 2022;47(1):62–79. doi:10.1080/17538157.2021.1929998

26. Shin HD, Durocher K, Lo B, Chen S, Ma C, Wiljer D, et al. Impact of a mental health patient portal on patients’ views of compassion: A mixed-methods study. BMC Digit Health. 2023;1(1). doi:10.1186/s44247-022-00002-z

27. Durocher K, Shin HD, Lo B, Chen S, Ma C, Strudwick G. Understanding the role of patient portals in fostering interprofessional collaboration within mental health care settings: Mixed methods study. JMIR Hum Factors. 2023;10:e44747. doi:10.2196/44747

28. Carini E, Villani L, Pezzullo AM, Gentili A, Barbara A, Ricciardi W, et al. The impact of digital patient portals on health outcomes, system efficiency, and patient attitudes: Updated systematic literature review. J Med Internet Res. 2021;23(9):e26189. doi:10.2196/26189

29. Norouzi Aval R, Rafatpanah H, Sarbaz M, Mousavi Baigi SF, Kimiafar K. Identification and classification features of patient portal, a systematic review. Health Sci Rep. 2025;8(3):e70520. doi:10.1002/hsr2.70520

